# Immediate effect of osteopathic techniques on human resting muscle tone in healthy subjects using myotonometry: A factorial randomized trial

**DOI:** 10.1101/2022.04.06.22273304

**Authors:** Lucas Bohlen, Jonah Schwarze, Jannik Richter, Bernadette Gietl, Christian Lazarov, Anna Kopyakova, Andreas Brandl, Tobias Schmidt

**Author notes:** Corresponding author: Name: Lucas Bohlen; Postal code: Mexikoring 19, 22297 Hamburg, Germany;.

## Abstract

**Background:** Musculoskeletal disorders (MSDs) are highly prevalent, burdensome, and putatively associated with an altered human resting muscle tone (HRMT). Osteopathic manipulative treatment (OMT) is commonly and effectively applied to treat MSDs and reputedly influences the HRMT. Arguably, OMT may modulate alterations in HRMT underlying MSDs. However, there is sparse evidence even for the effect of OMT on HRMT in healthy subjects.

**Methods:** A 3x3 factorial randomised trial was performed to investigate the immediate-term effect of myofascial release (MRT), muscle energy (MET), and soft tissue techniques (STT) on the HRMT of the corrugator supercilii (CS), superficial masseter (SM), and upper trapezius muscles (UT) in healthy subjects in Hamburg, Germany. Participants were randomised into three groups (1:1:1 allocation ratio) receiving treatment, according to different muscle-technique pairings, over the course of three sessions with one-week washout periods. Primarily, we assessed the effect of osteopathic techniques on muscle tone (F), biomechanical (S, D), and viscoelastic properties (R, C) from baseline to follow-up (main effect) and tested if specific muscle-technique pairs modulate the effect pre- to post-intervention (interaction effect) using the MyotonPRO (at rest). Data were analysed using descriptive (mean, standard deviation, quantiles, and simple effect) and inductive statistics (Bayesian ANOVA).

**Results:** 59 healthy participants were randomised into three groups and two subjects dropped out from one group (n=20; n=20; n=19 and n=17, respectively). The CS produced frequent measurement errors and was excluded from analysis. The main effect changed significantly for F (-0.163 [0.060]; p=0.008), S (-3.060 [1.563]; p=0.048), R (0.594 [0.141]; p<0.001), and C (0.038 [0.017]; p=0.028) but not for D (0.011 [0.017]; p=0.527). The interaction effect did not change significantly (p>0.05). No adverse events were reported.

**Conclusion:** OMT modified the HRMT in healthy subjects which may inform future research on MSDs. In detail, MRT, MET, and STT reduced the muscle tone (F), decreased biomechanical (S not D), and increased viscoelastic properties (R and C) of the SM and UT (CS was not measurable) at immediate term. However, the effect on HRMT was not modulated by muscle–technique interaction.

**Trial registration:** German Clinical Trial Register (DRKS00020393).

## INTRODUCTION

### Background

Globally, musculoskeletal disorders (MSDs) accounted for ∼1.3 billion prevalent and ∼334.7 million incident cases in 2017 [1]. Notably, most of the prevalence and incidence is attributable to gout, rheumatoid arthritis, osteoarthritis, neck pain (NP), and low back pain (LBP) [2, 3]. In 2017, MSDs were the main contributor to global disability and LBP was the leading cause of disability since 1990 [4]. Similarly, the global costs of MSDs due to health expenditure and production loss are reported to be immense [5]. However, these high health costs mismatch with low research investments [6] and policy responses are thus required to close the gap [7]. Hence, MSDs are highly prevalent, burdensome, and costly.

In primary care, musculoskeletal pain should be managed according to current best evidence and practice recommendations, which report beneficial effects of manual therapy but merely endorse adjunct treatment [8] due to limited high-quality evidence [9]. Still, patients with MSDs like LBP seem to prefer non-surgical [10] and non-pharmacological interventions [11] of whom manual therapy provides the best evidence for immediate-term reduction of pain and disability in acute and subacute non-specific LBP [12]. Hence, patients with MSDs may consult an osteopath in primary care – depending on varying country regulations and professional recognitions across the world [9, 13]. Osteopathic practice should, in turn, adhere to established treatment recommendations, which it arguably does **(Box 1)**.

#### Box 1 Osteopathic practice and musculoskeletal care recommendations

A recent systematic review of high-quality clinical practice guidelines put forward 11 recommendations for best practice care in musculoskeletal pain conditions emphasising: (1) patient centred care, (2) screening for red flags, (3) assessing of psychosocial factors, (4) using imaging only selectively, (5) carrying out a physical examination, (6) recording the patients’ progress, (7) providing patient education, (8) fostering physical activity and exercise, (9) applying manual therapy in multimodal care setting, (10) offering non-surgical care first, and (11) keeping patients working [8]. Arguably, osteopathy fulfils most of the criteria by: (1) being a patient-centered, or even person-centered, care approach [14, 15], (2) using an initial biomedical screening for red flags/serious underlying pathology [16], (3) assessing, and putatively influencing, psychosocial factors [17, 18], (4) using imaging only occasionally and mostly by referral [19, 20], (5) always carrying out physical examinations [21], (6) keeping patient records to monitor progress [20], (7) providing patient education [13, 22], (8) giving advice on physical activity and exercise [13, 23], (9) using manual therapy and recognizing the need for multimodal care [24], (10) offering non-surgical care, sometimes before and/or after surgery [25], and (11) aiming to promote the patient’s return to work [26, 27].

Osteopathy is a person-centered approach to healthcare that aims to enhance, restore, or maintain the patient’s (self-regulation of) structure, function, and well-being [13, 21]. Therefore, osteopaths employ both manual (e.g., touch, palpation, and manipulation) and patient management approaches (e.g., patient education, psychological support, lifestyle advice, and self-management solutions) [28]. In practice, palpatory findings are interpreted according to an osteopathic clinical reasoning process [16] that considers biopsychosocial perspectives [29] and osteopathic models of care [30]. These findings are then treated using osteopathic manipulative treatment (OMT) [31], which encompasses different approaches (e.g., direct, indirect, and combined methods) and techniques (e.g., myofascial release, muscle energy, and soft tissue techniques) [32] **(Box 2)**.

#### Box 2 Overview about myofascial release, muscle energy, and soft tissue techniques

Herein, we provide an overview about the definition, mechanisms, and effects of three manual techniques frequently used in the osteopathic field: **(1) Myofascial release technique (MRT)**: **(1.1) Definition**: MRT uses pressure and stretch with low load and long duration (which are adjusted based on palpatory feedback) to release myofascial tissues [33, 34]; **(1.2) Mechanisms**: MRT is underlined by mechanisms that are not fully understood [35, 36], however, it seems to induce fibroblasts to upregulate the production of anti-inflammatory cytokines and growth factors [37]; and **(1.3) Effects**: MRT shows mixed results for treating painful (often chronic) musculoskeletal conditions [33, 34, 38] as it reduces pain and improves function in patients with cLBP [39] while reducing disability, but not pain, in patients with LBP [40]; **(2) Muscle energy technique (MET)**: **(2.1) Definition**: MET involves instructing the patient to voluntarily contract muscles into a controlled direction against the therapist counter-pressure [41]; **(2.2) Mechanisms**: MET has unclear mechanisms of action with some speculating changes in proprioception, inflammation, and fluid circulation [42] and others emphasising post-isometric relaxation and reciprocal inhibition [43]; and **(2.3) Effects**: MET is effective in improving pain, disability, and range of motion in symptomatic and asymptomatic subjects, specifically in chronic MSDs like cLBP [43]; and **(3) Soft tissue technique (STT)**: **(3.1) Definition**: STT applies stretch, traction and/or deep pressure to soft tissues [32]; herein, we used repeated, slow, and deep pressure gliding strokes applied with the thumb alongside the muscle fibres, which is a STT that is similar to muscle stripping massage [44–46]; **(3.2) Mechanisms**: STT (or therapeutic massage, respectively) might work through mechanotransduction [47, 48] albeit other biomechanical, physiological, neurological, and psychological mechanisms are not to be disregarded [49, 50]; and **(3.3) Effects**: STT (or massage therapy, respectively) benefits patients with LBP [51, 52] and improves pain and function in patients with cLBP in the short-term [50].

OMT is primarily, but not exclusively, applied to treat musculoskeletal pain conditions like back pain [53] employing, among others, spinal [54] and visceral manipulations [55]. Current evidence suggests that osteopathic treatment may improve pain and function in patients with spinal complaints [56], including chronic NP [57] and acute and chronic non-specific LBP [58–60] even during pregnancy and postpartum [61]. OMT was recommended for patients with LBP [31], indicated to benefit medical care [62], proposed to be included within chronic pain management guidelines [63], and even reported to be dominant and cost-effective compared to usual care in the management of LBP and NP, respectively [64]. Still, the current body of evidence lacks robustness due to methodological shortfalls and counterevidence is available as well [65–67].

Taken together, MSDs and OMT are complex health conditions and interventions, respectively. Both are underlined by mechanisms that are poorly understood [30, 68] and associated with various biological, psychological, and social factors [69, 70]. However, of particular interest is that both MSDs [71, 72] and OMT [73–76] are reputed to be associated with changes in muscle tone. On the one hand, alterations in lumbar myofascial tone and stiffness seem to contribute to the development of LBP [77] and may be linked to underlying pathologies and symptoms [71, 72]. On the other hand, manual osteopathic treatment is assumed to alter muscle tone and stiffness [73–76], for example, of paraspinal muscles in patients with LBP [71]. Therefore, we hypothesise that a putative mechanism of action underpinning the treatment of MSDs with OMT might be the modulation of muscle tone.

In general, muscle tone relates to the resting tension of the tissue in response to stretch. A differentiation can be drawn between: (1) active muscle tone: relating to the electrical activity within muscle cells; and (2) passive muscle tone: relating to the intrinsic biomechanical and viscoelastic properties of the muscle [72, 78]. The human resting muscle tone (HRMT) broadens the concept of passive muscle tone to include other connective tissues thereby accounting for the passive/resting tension of all continuous myofascial tissues involved in the body’s biotensegrity system [79]. It was suggested that the HRMT is generated by ‘slowly cycling cross-bridges’ between myosin heads and actin filaments [77, 79, 80]; however, other molecular and cellular mechanisms have been put forward and further research is required to conclusively determine the mechanisms underlying the HRMT [81].

Notably, the HRMT seems to be particularly relevant to clinical and manual practice [79]. Still, previous studies assessing the presumed effect of osteopathy on muscle tone used palpation and electromyography (EMG) as measures [82–84]. However, manual palpation is reported to be unreliable [85–92] and EMG is not informative of the HRMT [79]. Nonetheless, it appears imperative to detect these changes in research and practice. Thus, the use of a myotonometer was emphasized to reduce the subjectivity in determining the HRMT [71, 81]. An objective and reliable myotonometer is the MyotonPRO which induces oscillations in the muscle fibres as a means of quantifying biomechanical and viscoelastic muscle properties [71, 72, 93].

### Objectives

Hence, in this study, we assessed the effect of osteopathic modalities on the HRMT in healthy subjects using the MyotonPRO to inform future research on MSDs. In detail, three osteopathic techniques with different characteristics (myofascial release technique [MRT], muscle energy technique [MET], and soft tissue technique [STT]) were used to influence the HRMT of three muscles with different sizes and thicknesses (corrugator supercilii muscle [CS], superficial masseter muscle [SM], and upper trapezius muscle [UT]) **(Box 3)**. Primarily, we evaluated whether osteopathic techniques influence the tone (F), biomechanical (S and D) and viscoelastic (R and C) properties of these muscles in all groups from baseline to follow-up using the MyotonPRO (main effect). Secondarily, we investigated if muscle-technique pairs modulate the putative effect in each group from pre-to post-intervention using the MyotonPRO (interaction effect). For the primary objective, we hypothesized that osteopathic techniques reduce the muscle tone (F) and biomechanical properties (S and D) and increase the viscoelastic properties (R and C). For the secondary objective, we hypothesized that the predicted changes in muscle properties of the CS, SM, and UT are preferentially achieved through MRT, MET, and STT, respectively.

#### Box 3 Muscle characteristics

The CS, SM, and UT were selected for this study due to their apparent differences in size and thickness. However, the availability of data on both size and thickness were scattered and the values given below are to be interpreted with caution. In detail, it has been reported that the relaxed muscle thickness of the (1) CS ranges between 5 and 6 mm [94]; (2) SM ranges between 9 and 15 mm (notably, values account for the combined thickness of the superficial and deep part of the masseter muscle and are thus exaggerated) [95], and (3) UT ranges between 11 and 12 mm [96]. No data was available on the surface size of these muscles. Hence, we calculated an approximate surface size based on data reporting the length and width of these muscles (which is imprecise as it does not account for factors like muscle shape). In detail, we calculated a surface size for the (1) CS of ∼3.69 cm^2^ (length: 29.24 mm; width: 12.62 mm) [97]; (2) SM of ∼24.40 cm^2^ (length: 6.32 cm; width: 3.86 cm) [98]; and (3) UT of ∼540 cm^2^ (length: 45 cm; width: 12 cm) (notably, this data is based on a myocutaneous trapezius flap and likely imprecise) [99].

## METHODS

### Trial design

This single-blinded 3x3 factorial randomised trial was conducted in Hamburg, Germany. No changes to the methods were made after trial commencement. The study is largely reported according to the CONSORT statement [100, 101] since there are currently no specific guidelines available for randomised trials using a factorial design [102]. This study has obtained informed consent from participants, was carried out in accordance with the Declaration of Helsinki [103] and was approved by the ethics committee of the Osteopathic Research Institute (Nr. 020-01). The study was prospectively registered within the German Clinical Trial Register (DRKS00020393).

### Trial procedure

Participants were randomly allocated into three groups (G1, G2, and G3) undergoing three treatment periods (t1-t3). During each period, each group received treatment with the same osteopathic technique but applied by another practitioner to another muscle. Over the course of the trial, all three groups were treated (1) with all three osteopathic techniques (MRT, MET, and SST); (2) at all three muscles (CS, SM, and UT); and (3) by all three practitioners (P1, P2, P3). However, the muscle-technique-practitioner combination was distinct for each group during each period **(Table 1)**. All interventions and measurements were applied (1) to the right side of the participant’s body to ensure comparability, and (2) in relaxed supine position to maintain resting muscle state. The trial comprised one-week washout periods between baseline (t0) and each session (t1-t3). A session consisted of one intervention day which encompassed 5 minutes of measurement, followed by 5 minutes of treatment, and renewed 5 minutes of measurement per subject. Participants started with 5 minutes delay to one another to allow measurement by one assessor who was not involved with the interventions. In this trial, all groups were intervention groups that were treated at one muscle per session, while all three muscles were measured. Thus, measures from untreated muscles were used as control values **(Table 2)**.

**Table 1.**
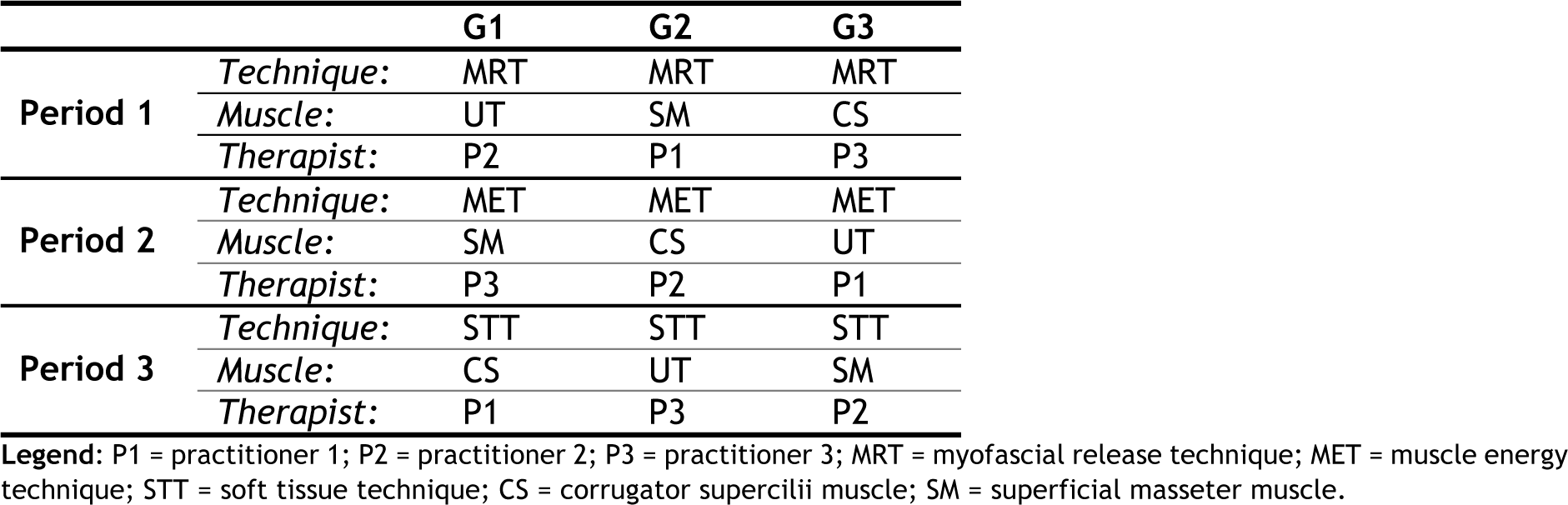
Trial procedure

|  |  | G1 | G2 | G3 |
| --- | --- | --- | --- | --- |
| Period 1 | *Technique:* | MRT | MRT | MRT |
|  | *Muscle:* | UT | SM | CS |
|  | *Therapist:* | P2 | P1 | P3 |
| Period 2 | *Technique:* | MET | MET | MET |
|  | *Muscle:* | SM | CS | UT |
|  | *Therapist:* | P3 | P2 | P1 |
| Period 3 | *Technique:* | STT | STT | STT |
|  | *Muscle:* | CS | UT | SM |
|  | *Therapist:* | P1 | P3 | P2 |

Legend: P1 = practitioner 1; P2 = practitioner 2; P3 = practitioner 3; MRT = myofascial release technique; MET = muscle energy technique; STT = soft tissue technique; CS = corrugator supercilii muscle; SM = superficial masseter muscle.

**Table 2.** Controls.

|  |  |  |
| --- | --- | --- |
| Period 1 | G1 | CS (control) |
|  |  | SM (control) |
|  |  | UT (intervention) |
|  | G2 | CS (control) |
|  |  | SM (intervention) |
|  |  | UT (control) |
|  | G3 | CS (intervention) |
|  |  | SM (control) |
|  |  | UT (control) |
| Period 2 | G1 | CS (control) |
|  |  | SM (intervention) |
|  |  | UT (control) |
|  | G2 | CS (intervention) |
|  |  | SM (control) |
|  |  | UT (control) |
|  | G3 | CS (control) |
|  |  | SM (control) |
|  |  | UT (intervention) |
| Period 3 | G1 | CS (intervention) |
|  |  | SM (control) |
|  |  | UT (control) |
|  | G2 | CS (control) |
|  |  | SM (control) |
|  |  | UT (intervention) |
|  | G3 | CS (control) |
|  |  | SM (intervention) |
|  |  | UT (control) |

### Participants

Three undergraduate classes of osteopathy students were recruited from the Osteopathie Schule Deutschland in Hamburg, Germany. The sample was limited to healthy subjects from this specific setting. The eligibility criteria were specified to include participants between 18 and 50 years old and exclude participants with health complaints (particularly muscle disorders) to minimise the risk of age- [104, 105] and disease-related [106] changes of the musculature.

### Interventions

Three manual techniques from the osteopathic field (MRT, MET, and STT) were selected and administered for ∼5 minutes. These osteopathic techniques were applied with the aim of modifying the HRMT in healthy participants and were adjusted to fit the structure and function of each muscle. A consensus training was implemented prior to the trial to ensure that all therapists applied the interventions coherently. During the first session, MRT was applied to the right CS (G3), SM (G2), and UT (G1). During the second session, MET was applied to the right CS (G2), SM (G1), and UT (G3). During the third session, STT was applied to the right CS (G1), SM (G3), and UT (G2) **(Table 1)**. Overall, the interventions were chosen to represent the broad range of osteopathic techniques and their diverse characteristics comprising: (1) direct, indirect, and combined techniques; (2) active and passive techniques; and (3) techniques with high- and low-pressure or counterforce [32, 107].

#### Myofascial Release Technique (MRT)

MRT is an indirect (or direct) and passive technique using low pressure. The muscle is palpated (covering origin and insertion) and guided alongside the path of least resistance into a position of ease [108], thereby following the tissues’ micro– movements away from the restricted barrier until a release occurs (**Figure 1**).

**Figure 1.** Osteopathic techniques; **Legend**: red arrows = therapists’ motion; black arrows = participants’ motion; four-headed arrows = motion applied in all directions.

#### Muscle Energy Technique (MET)

MET is a direct and active technique using counterforce. The therapist brings the muscle into a position of stretch and holds it at the restriction barrier. The participant then performs an isometric contraction of the muscle (with 25% of maximum effort/force) away from this restricted barrier and against the therapist’s counterforce [109]. After approximately 3–6 seconds of contraction, the participant relaxes, and the therapist adjusts the tissue towards its renewed movement/restriction barrier while following the tissue’s micro–movements. This post–isometric relaxation approach is repeated 3–6 times (**Figure 1**).

#### Soft Tissue Technique (STT)

STT is a direct and passive technique using high pressure. The therapist applies repeated longitudinal deep pressure gliding strokes with the thumb alongside the muscle fibres of the CS (from origin towards insertion), SM (from origin towards insertion), and UT (from insertion towards origin). This is similar to the treatment of a trigger band according to the fascial distortion model [110] but is applied to palpably firm muscle fibres and their fascial surroundings (**Figure 1**).

### Outcomes

We used the handheld digital palpation device MyotonPRO [Version 5.0.0] as the outcome measure (**Figure 2**). This myotonometer assesses the muscle’s tone, biomechanical and viscoelastic properties using five parameters by means of dynamic oscillation mechanosignals [81, 111] **(Table 3)**. The MyotonPRO is a valid and reliable measurement tool for healthy and diseased participants [112, 113] **(Box 4)** that has been applied to evaluate muscle tone, muscle stiffness, and HRMT in multiple studies investigating various structures and conditions [71, 81, 93, 114– 116]. The myotonometer measurements were carried out at all three sessions (t1-t3) before and after the treatment intervention. Measurement points (MPs) were predefined for the myotonometer measurements of each muscle prior to the trial **(Box 5)**. MPs were identified by manual palpation following anatomical landmarks. All MPs were marked before each session using a dermatological skin marker pen.

**Figure 2.** MyotonPRO; **Legend**: Not applicable.

**Table 3.** MyotonPRO parameters.

| Parameter | Description | Formula |
| --- | --- | --- |
| F - Oscillation Frequency | Measures the muscle's tone in resting state (excluding voluntary contractions like depicted within an EMG measurement) in Hertz (Hz) | $F = f_{max}$ |
| S - Dynamic Stiffness | Measures the biomechanical property of a muscle to deform its shape under internal or external force in Newton meter (N/m) | $S = \frac{a_{max} \cdot m_{probe}}{\Delta l}$ |
| D - Logarithmic Decrement | Measures the elasticity characterized by the muscle's natural oscillation (which represents the biomechanical ability to regain its initial shape after deformation under internal or external force) | $D = \ln\left(\frac{a_1}{a_3}\right)$ |
| R - Mechanical Stress Relaxation Time | Measures the time the muscle tissue needs to recover its shape after deformation under internal or external force in milliseconds (ms) | $R = t_R - t_1$ |
| C - Ratio of deformation and Relaxation time | Measures the gradual elongation of muscle tissue over time under constant tensile stress | $C = \frac{R}{t_1 - t_T}$ |

#### Box 4 Validity and reliability of the MyotonPRO

The MyotonPRO shows good validity and high reliability for measuring, for example, the trapezius muscle [117]. In detail, studies demonstrate moderate to excellent inter-rater reliability (intra-class correlation coefficient [ICC] for F = 0.87, S = 0.79, D = 0.93, R = 0.65, C = 0.50; standard error of measurement [SEM] for F = 0.7, S = 16.8, D = 0.2, R = 1.4, C = 0.1), moderate to good intra-rater reliability (ICC for F = 0.81, S = 0.82, D = 0.76, R = 0.74, C = 0.52; SEM for F = 0.8, S = 16.9, D = 0.2, R = 1.2, C = 0.1) [118], and good to excellent test-retest reliability (ICC for S = 0.821-0.913; SEM for S = 23.59) [119].

#### Box 5 Measurement points (MPs)

The MP for the: (1) CS was determined to be located 0.5 cm superior to the supraorbital notch slightly above the eyebrow [120, 121]; (2) SM was determined to be located just below the midpoint of a virtual line between the muscle’s origin and attachment (masseteric tuberosity of the mandibular angle and the tendinous aponeurosis at the anterior third of the zygomatic arch) [122, 123]; and (3) UT was determined to be located halfway between a virtual line from the top of the acromion to the spinous process of C7 (which is ∼ 19.5 cm) [81, 124] (notably, MPs were inspired, not determined, by the cited references).

### Sample size

The sample size was calculated prospectively using G*Power, which is a power analysis for ANOVA with repeated measures (within-between interaction) [125, 126]. We assumed a type I error level of 0.05 and statistical power of 95%. Based on an estimated partial ⴄ^2^ of 0.1 (unpublished data), an effect size of 0.33, and three measurements and four groups, a total sample size of 52 participants was calculated. Using an estimated drop-out rate of 15%, the sample size was planned with 60 participants.

### Randomisation

The sample was randomly allocated into three groups (G1, G2, G3) by block randomization (1:1:1 allocation ratio) using computer-generated allocation schedule (http://www.randomization.com). Furthermore, we randomly assigned which technique would be applied in which period by throwing the dice. Afterwards, we randomly assigned the muscles and therapists to the groups and periods in the same manner. The principal investigator generated the random allocation sequence, enrolled participants, and assigned them to sequences of intervention while having no clinical involvement in the trial. Treatments were scheduled according to allocation sequence and therapists and participants were first introduced to each other during the respective sessions.

### Blinding

Participants and statisticians, but not therapists and assessors, were blinded to the conditions. However, we assume that blinding was compromised because the participants were osteopathy students that were likely able to distinguish between the interventions.

### Statistical methods

The outcomes from myotonometer measurements were assessed by calculating the within-participant difference of each parameter for all groups between t1 and t3 (primary objective) and the between-participant difference of each parameter for each group between pre- and post-treatment of each session (secondary objective). Statistical analysis was conducted by employing the software R Studio. Myotonometer properties (F, S, D, R, and C) were used as parameters and converted into factors. Heidelberger & Welch’s diagnostic was used to run length diagnostic and convergence diagnostic. The descriptive statistic for the primary objective was presented by mean, standard deviation, and quantiles, whereas the secondary objective was presented by standard deviation, quantiles, and simple effect. Due to the limitations of standard repeated measures ANOVA for categorical variables and unbalanced data, the inductive statistics for the primary and secondary objectives as well as subgroup analysis were calculated using the Bayesian version of the repeated-measures analysis of variance (BANOVA) [127]. The p-values of the multiple comparisons were adjusted using the Bayesian model [128]. The significance level was set to 5% (p≤0.05). Missing completely at random values were included for further analysis. Missing at random and missing not at random values were excluded as they are dependent on one factor and bias the results [129].

## RESULTS

Overall, 82 participants were screened and 23 declined to participate or had scheduling issues. The remaining 59 participants were randomly allocated into groups, leading to a sample of 20 participants for groups G1 and G2, and 19 participants for the G3 group. Two participants from group G3 cancelled their participation in the study due to personal reasons and were excluded from interventions and analysis prior to the first period. Therefore, the first trial period started with the final sample size (n=57) distributed across groups as follows: G1 (n=20), G2 (n=20), and G3 (n=17). Unfortunately, some participants missed scheduled appointments and thereby allocated interventions and measurements. These participants were not excluded from study participation and data was included for analysis unless all three periods were missed, which did not occur. In detail, appointments were missed during the: (1) first period by two participants from group G1 and one participant from group G2; (2) second period by three participants from group G3; and (3) third period by five participants from group G1, two participants from group G2, and two participants from group G3 (**Figure 3**).

**Figure 3.**
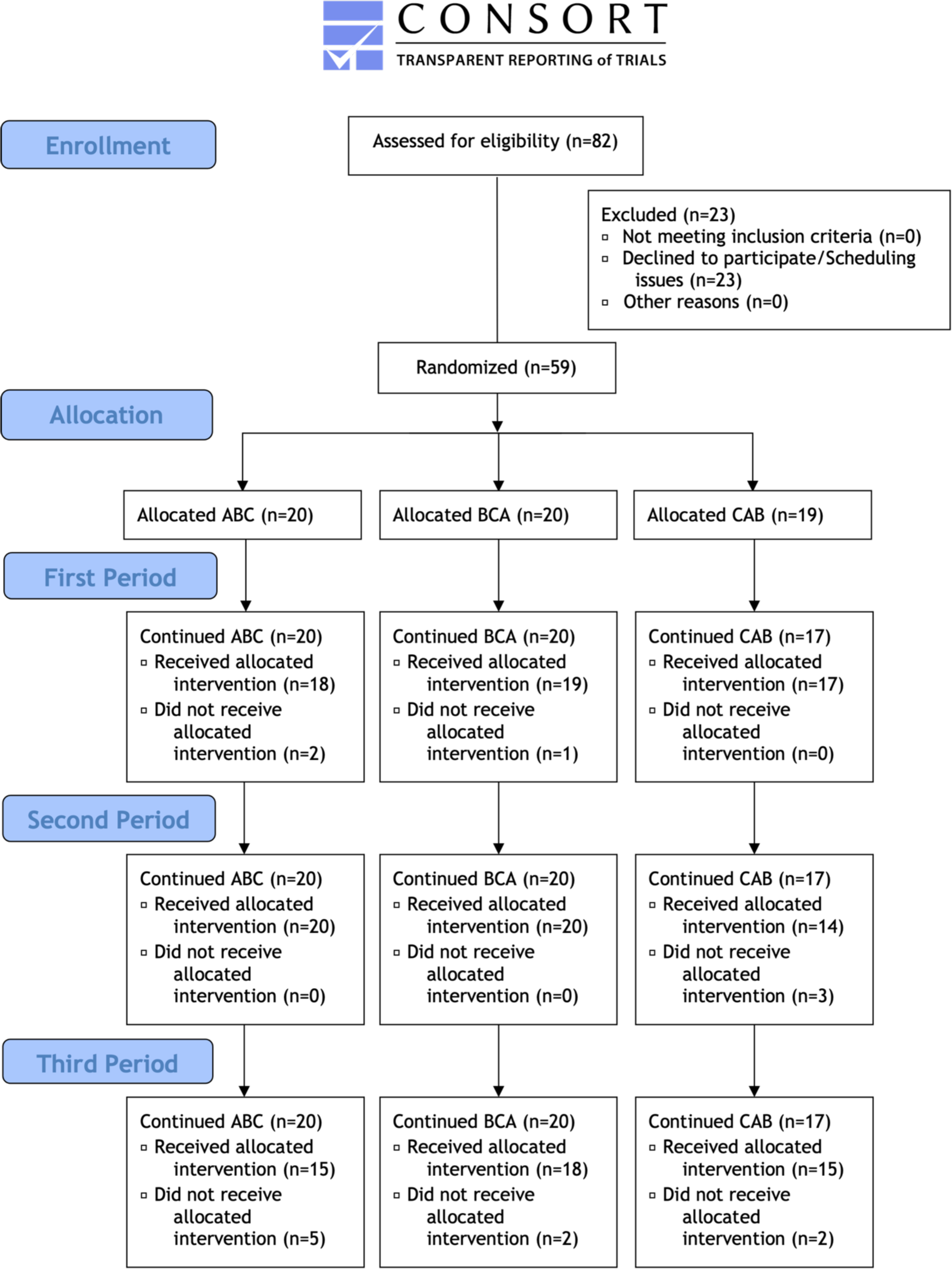
Participant flow chart; **Legend**: Not applicable.

### Recruitment

Participants were recruited, provided written informed consent, and reported baseline data in December 2019. The trial was implemented over the course of one month between January and February 2020.

### Baseline data

Baseline demographics were recorded and included sex, handedness, age, and body mass index (BMI). The sample was predominantly female (68%), right-handed (91%), young (22.7 ± 4.5 years), and of normal weight (22.0 ± 2.5 BMI) **(Table 4)**.

**Table 4.** Baseline demographics.

| Parameter | G1 (n=20) |  | G2 (n=20) |  | G3 (n=17) |  |
| --- | --- | --- | --- | --- | --- | --- |
|  | *R* | % | *R* | % | *R* | % |
| F:M | 14:6 | 70 | 13:7 | 65 | 12:5 | 71 |
| RH:LH | 16:4 | 80 | 19:1 | 95 | 17:0 | 100 |
|  | *Mean* | *SD* | *Mean* | *SD* | *Mean* | *SD* |
| Age | 22.0 | 1.7 | 21.4 | 2.7 | 25.1 | 7.0 |
| BMI | 21.8 | 1.7 | 21.3 | 2.1 | 23.2 | 3.4 |

Legend: F = female; M = male; RH = right-handed; LH = left-handed; R = ratio; SD = standard deviation.

### Numbers analysed

The data were examined for availability and normality to rule out statistical errors during analysis. Overall, 8.9% of the collected data did not correspond to the confidence interval of the MyotonPRO (set to 90%) and was thus not available for analysis. All missing values arose from measurements of the CS, demonstrating that the data was not missing at random. The CS was consequently excluded from analysis because correct coefficient estimation was not guaranteed. After exclusion, data from 104 treatments (54 for SM and 50 for UT) were included for analysis (104 of 156 measures).

### Outcomes

Here, full outcome data are reported **(Table 5)**. Subsequently, results will be presented (excluding the CS) according to the primary and secondary objectives.

**Table 5.** Results – Outcomes.

|  |  |  |  | F | S | D | R | C | F | S | D | R | C | F | S | D | R | C |
| --- | --- | --- | --- | --- | --- | --- | --- | --- | --- | --- | --- | --- | --- | --- | --- | --- | --- | --- |
| MRT | CS | pre | mean | 31.24 | 1398.50 | 1.79 | 6.03 | 0.62 | 32.17 | 1565.75 | 1.72 | 5.70 | 0.60 | 32.09 | 1445.75 | 1.70 | 5.87 | 0.64 |
|  |  |  | SD | 2.61 | 224.15 | 0.12 | 0.73 | 0.52 | 4.96 | 35.09 | 0.14 | 0.83 | 0.05 | 4.24 | 142.66 | 0.11 | 0.65 | 0.07 |
|  |  | post | mean | 32.73 | 1301.67 | 1.72 | 6.18 | 0.61 | 32.14 | 1352.83 | 1.74 | 6.07 | 0.61 | 33.35 | 1367.63 | 1.75 | 6.02 | 0.62 |
|  |  |  | SD | 3.85 | 293.65 | 0.11 | 1.15 | 0.07 | 3.25 | 197.70 | 0.11 | 0.83 | 0.06 | 3.43 | 179.33 | 0.11 | 0.89 | 0.06 |
|  | SM | pre | mean | 14.24 | 265.11 | 1.60 | 22.65 | 1.40 | 13.90 | 270.47 | 1.67 | 22.22 | 1.38 | 14.09 | 264.44 | 1.62 | 22.72 | 1.41 |
|  |  |  | SD | 1.73 | 49.29 | 0.23 | 3.20 | 0.16 | 1.36 | 34.34 | 0.16 | 2.63 | 0.15 | 1.24 | 39.49 | 0.21 | 3.21 | 0.18 |
|  |  | post | mean | 14.08 | 262.78 | 1.59 | 22.96 | 1.41 | 13.79 | 257.40 | 1.62 | 22.97 | 1.42 | 13.97 | 260 | 1.59 | 23.01 | 1.43 |
|  |  |  | SD | 1.49 | 47.98 | 0.26 | 2.55 | 0.13 | 0.97 | 32.10 | 0.19 | 2.76 | 0.16 | 1.23 | 34.67 | 0.18 | 3.06 | 0.18 |
|  | UT | pre | mean | 12.11 | 193.53 | 1.16 | 26.48 | 1.56 | 11.72 | 167.73 | 0.99 | 27.44 | 1.56 | 11.68 | 171.06 | 1.01 | 27.52 | 1.56 |
|  |  |  | SD | 0.97 | 30.18 | 0.25 | 2.60 | 0.15 | 0.64 | 20.28 | 0.12 | 2.39 | 0.19 | 0.93 | 33.65 | 0.18 | 1.93 | 0.10 |
|  |  | post | mean | 11.93 | 184.29 | 1.10 | 26.61 | 1.52 | 11.62 | 170.84 | 1.00 | 27.78 | 1.60 | 11.54 | 172.22 | 1.03 | 28.13 | 1.61 |
|  |  |  | SD | 0.88 | 37.03 | 0.21 | 2.62 | 0.20 | 0.69 | 21.53 | 0.11 | 2.16 | 0.13 | 0.87 | 29.76 | 0.15 | 2.13 | 0.09 |
| MET | CS | pre | mean | 33.37 | 1370.75 | 1.79 | 6.19 | 0.62 | 33.04 | 1376.33 | 1.82 | 6.28 | 0.62 | 31.56 | 1311 | 1.81 | 6.01 | 0.63 |
|  |  |  | SD | 3.11 | 193.16 | 0.09 | 0.68 | 0.04 | 3.85 | 169.39 | 0.08 | 1.03 | 0.06 | 2.96 | 20.04 | 0.11 | 0.64 | 0.05 |
|  |  | post | mean | 32.81 | 1325.40 | 1.74 | 6.39 | 0.64 | 32.67 | 1378.13 | 1.83 | 6.15 | 0.63 | 33.05 | 1504.50 | 1.80 | 6.28 | 0.63 |
|  |  |  | SD | 3.58 | 205.99 | 0.12 | 1.09 | 0.07 | 4.42 | 199.35 | 0.10 | 0.88 | 0.07 | 5.24 | 167.61 | 0.13 | 0.67 | 0.04 |
|  | SM | pre | mean | 13.98 | 283.85 | 1.79 | 20.93 | 1.30 | 13.79 | 257.40 | 1.62 | 22.97 | 1.42 | 13.96 | 267.50 | 1.67 | 22.95 | 1.44 |
|  |  |  | SD | 1.07 | 31.12 | 0.18 | 2.52 | 0.15 | 0.97 | 31.25 | 0.20 | 2.68 | 0.16 | 1.03 | 32.69 | 0.17 | 2.62 | 0.15 |
|  |  | post | mean | 13.95 | 283.15 | 1.75 | 21.19 | 1.32 | 11.75 | 252.35 | 1.61 | 23.86 | 1.48 | 14.13 | 270.71 | 1.65 | 22.52 | 1.40 |
|  |  |  | SD | 1.12 | 33.46 | 0.15 | 2.48 | 0.15 | 1.03 | 37.04 | 0.20 | 2.88 | 0.17 | 1.23 | 32.04 | 0.21 | 2.47 | 0.15 |
|  | UT | pre | mean | 11.71 | 170.45 | 1.01 | 27.41 | 1.55 | 11.51 | 171.95 | 1.03 | 28.09 | 1.60 | 11.84 | 182.21 | 1.08 | 27.45 | 1.60 |
|  |  |  | SD | 0.83 | 26.19 | 0.11 | 1.92 | 0.12 | 0.89 | 28.44 | 0.15 | 2.36 | 0.14 | 0.89 | 23.43 | 0.21 | 2.62 | 0.14 |
|  |  | post | mean | 11.55 | 169.05 | 1.01 | 27.95 | 1.58 | 11.31 | 168.6 | 1.05 | 28.60 | 1.62 | 11.75 | 181.93 | 1.13 | 27.61 | 1.60 |
|  |  |  | SD | 0.83 | 26.65 | 0.13 | 2.19 | 0.13 | 0.81 | 28.09 | 0.15 | 2.30 | 0.14 | 0.89 | 27.18 | 0.26 | 3.00 | 0.17 |
| STT | CS | pre | mean | 33.29 | 1475.50 | 1.77 | 6.02 | 0.61 | 35.25 | 1472.25 | 1.81 | 5.54 | 0.60 | 33.77 | 1387.86 | 1.75 | 6.22 | 0.62 |
|  |  |  | SD | 3.83 | 239.40 | 0.09 | 0.77 | 0.06 | 4.28 | 82.05 | 0.11 | 0.40 | 0.05 | 3.78 | 128.75 | 0.12 | 0.70 | 0.05 |
|  |  | post | mean | 33.61 | 1402.60 | 1.78 | 6.04 | 0.62 | 35.05 | 1428.50 | 1.77 | 5.84 | 0.61 | 34.39 | 1440.33 | 1.77 | 5.82 | 0.61 |
|  |  |  | SD | 4.45 | 140.40 | 0.07 | 0.64 | 0.04 | 5.17 | 228.56 | 0.10 | 0.75 | 0.04 | 4.12 | 196.24 | 0.10 | 0.65 | 0.04 |
|  | SM | pre | mean | 13.97 | 262.87 | 1.65 | 22.56 | 1.40 | 13.80 | 265.67 | 1.70 | 22.85 | 1.42 | 13.87 | 266.78 | 1.67 | 22.43 | 1.40 |
|  |  |  | SD | 0.89 | 29.16 | 0.24 | 2.52 | 0.15 | 1.08 | 26.10 | 0.19 | 2.10 | 0.13 | 1.24 | 27.23 | 0.15 | 2.44 | 0.14 |
|  |  | post | mean | 13.88 | 259.13 | 1.62 | 22.87 | 1.42 | 13.78 | 254.2 | 1.64 | 22.99 | 1.42 | 13.72 | 261.39 | 1.63 | 22.99 | 1.44 |
|  |  |  | SD | 0.90 | 31.03 | 0.23 | 2.77 | 0.16 | 1.23 | 30.57 | 0.22 | 2.42 | 0.13 | 1.11 | 27.31 | 0.15 | 2.47 | 0.14 |
|  | UT | pre | mean | 11.49 | 176.53 | 1.04 | 28.39 | 1.64 | 11.81 | 188.2 | 1.10 | 27.40 | 1.62 | 11.66 | 174.00 | 1.02 | 27.85 | 1.61 |
|  |  |  | SD | 0.97 | 26.21 | 0.15 | 3.27 | 0.20 | 0.53 | 21.09 | 0.23 | 1.53 | 0.09 | 0.83 | 17.21 | 0.11 | 2.02 | 0.14 |
|  |  | post | mean | 11.55 | 176.27 | 1.04 | 27.63 | 1.57 | 11.77 | 189.20 | 1.11 | 27.51 | 1.63 | 11.38 | 174.00 | 1.09 | 28.96 | 1.70 |
|  |  |  | SD | 1.00 | 35.47 | 0.17 | 2.80 | 0.18 | 0.53 | 21.23 | 0.19 | 1.59 | 0.10 | 0.68 | 12.62 | 0.15 | 1.87 | 0.10 |

Legend: yellow = G1; blue = G2; green = G3; darker colouring = treated muscles (intervention values); lighter colouring = untreated muscles (control values). Abbreviations: MRT = myofascial release technique; MET = muscle energy technique; STT = soft tissue technique; CS = corrugator supercilii muscle; SM = superficial masseter muscle; UT = upper trapezius muscle; pre = before intervention; post = after intervention; SD = standard deviation; F = Oscillation Frequency; S = Dynamic Stiffness; D = Logarithmic Decrement; R = Mechanical Stress Relaxation Time; C = Ratio of deformation and Relaxation time.

#### Outcomes for the primary objective

The outcomes for the primary objective were assessed by means of the standardized mean difference. The data passed all of Heidelberger’s and Welch’s convergence diagnoses and showed that: F (-0.163 [0.060]; p=0.008), S (-3.060 [1.563]; p=0.048), R (0.594 [0.141]; p<0.001), and C (0.038 [0.017]; p=0.028) changed significantly, while D (0.011 [0.017]; p=0.527) did not change significantly **(Table 6)**. In other words, muscle tone (F [p=0.008]) and biomechanical properties (S [p=0.048] not D [p=0.527]) decreased, while the viscoelastic properties (R [p<0.001] and C [p=0.028]) increased. Subgroup analysis for sex-specific changes revealed a significant interaction for F (-0.192 [0.089]; p=0.030), but not for S (0.008 [0.0125]; p=0.510), D (0.008 [0.013]; p=0.555), R (-0.423 [0.218]; p=0.057), and C (-0.019 [0.015]; p=0.237) **(Table 6)**.

**Table 6.**
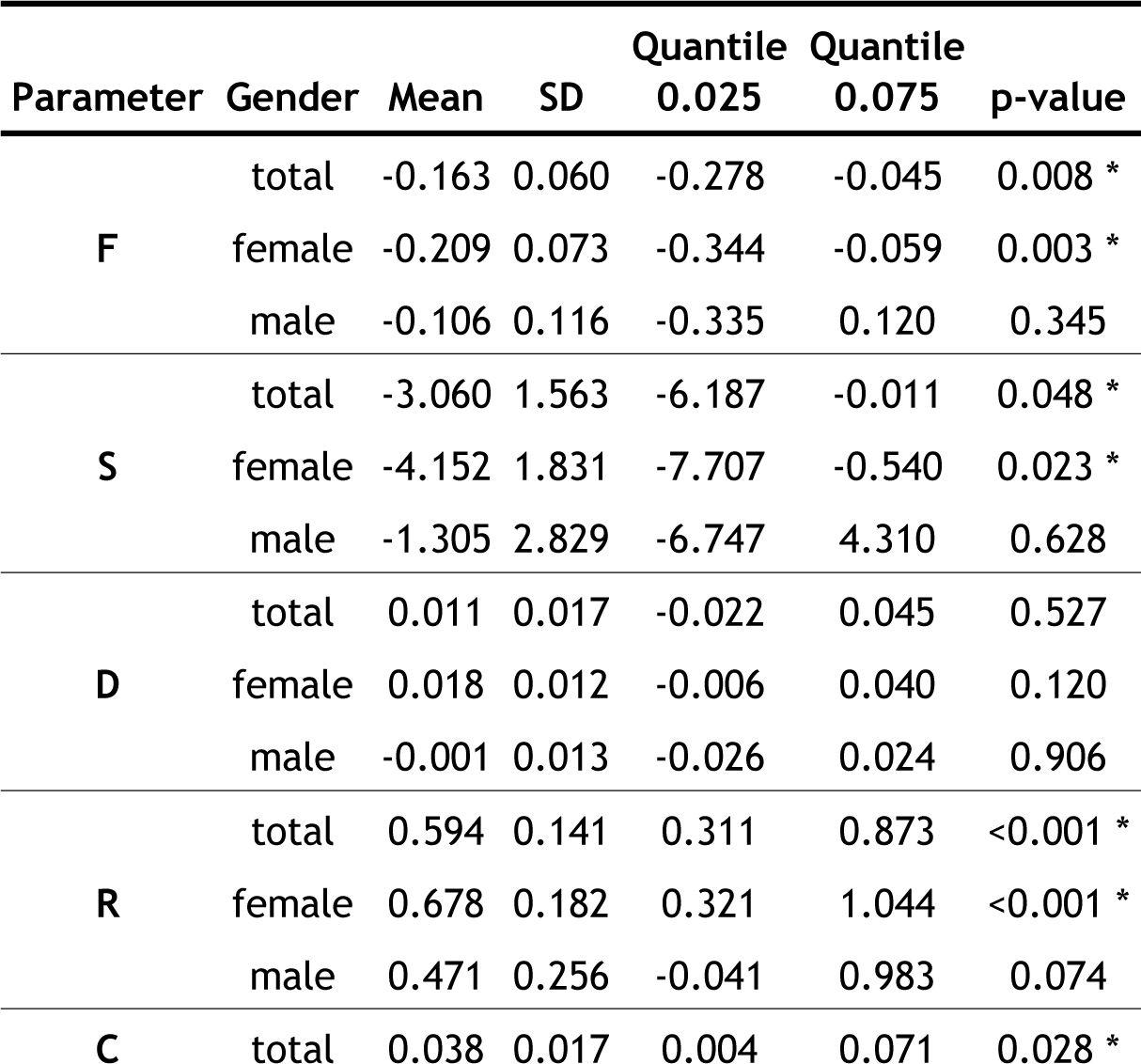
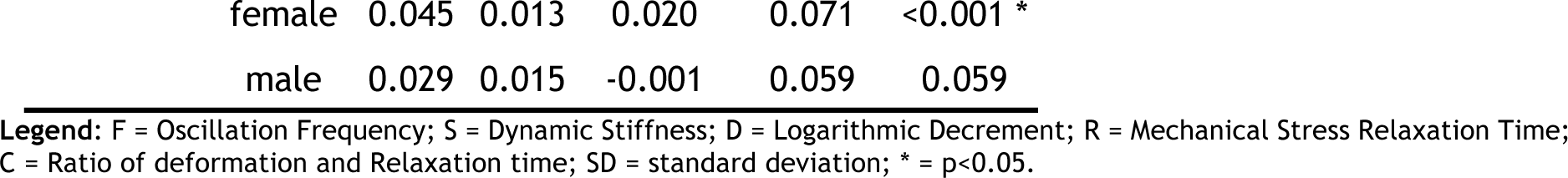
Results – Outcomes for the primary objective.

| Parameter | Gender | Mean | SD | Quantile<br>0.025 | Quantile<br>0.075 | p-value |
| --- | --- | --- | --- | --- | --- | --- |
| F | total | -0.163 | 0.060 | -0.278 | -0.045 | 0.008 * |
|  | female | -0.209 | 0.073 | -0.344 | -0.059 | 0.003 * |
|  | male | -0.106 | 0.116 | -0.335 | 0.120 | 0.345 |
| S | total | -3.060 | 1.563 | -6.187 | -0.011 | 0.048 * |
|  | female | -4.152 | 1.831 | -7.707 | -0.540 | 0.023 * |
|  | male | -1.305 | 2.829 | -6.747 | 4.310 | 0.628 |
| D | total | 0.011 | 0.017 | -0.022 | 0.045 | 0.527 |
|  | female | 0.018 | 0.012 | -0.006 | 0.040 | 0.120 |
|  | male | -0.001 | 0.013 | -0.026 | 0.024 | 0.906 |
| R | total | 0.594 | 0.141 | 0.311 | 0.873 | <0.001 * |
|  | female | 0.678 | 0.182 | 0.321 | 1.044 | <0.001 * |
|  | male | 0.471 | 0.256 | -0.041 | 0.983 | 0.074 |
| C | total | 0.038 | 0.017 | 0.004 | 0.071 | 0.028 * |

|  |  |  |  |  |  |
| --- | --- | --- | --- | --- | --- |
| female | 0.045 | 0.013 | 0.020 | 0.071 | <0.001 * |
| male | 0.029 | 0.015 | -0.001 | 0.059 | 0.059 |

Legend: F = Oscillation Frequency; S = Dynamic Stiffness; D = Logarithmic Decrement; R = Mechanical Stress Relaxation Time; C = Ratio of deformation and Relaxation time; SD = standard deviation; \* = $p < 0.05$ .

#### Outcomes for the secondary objective

Since there was a significant interaction between treatment and muscle (0.037 [0.014]; p=0.009), the simple effects were interpreted for the secondary objective. There was a tendency for a difference in comparison between MRI, MET and STT, but all multiple comparisons between treatment and muscle were not significant (p>0.05) **(Table 7)**. There was no sex-specific simple effect for the significant interaction but some tendency (p>0.05). For example, in males compared to females, F of the UT showed a higher increase and decrease following MET and MRT, respectively **(Table 8) (Figure 4) (Figure 5)**.

**Figure 4.**
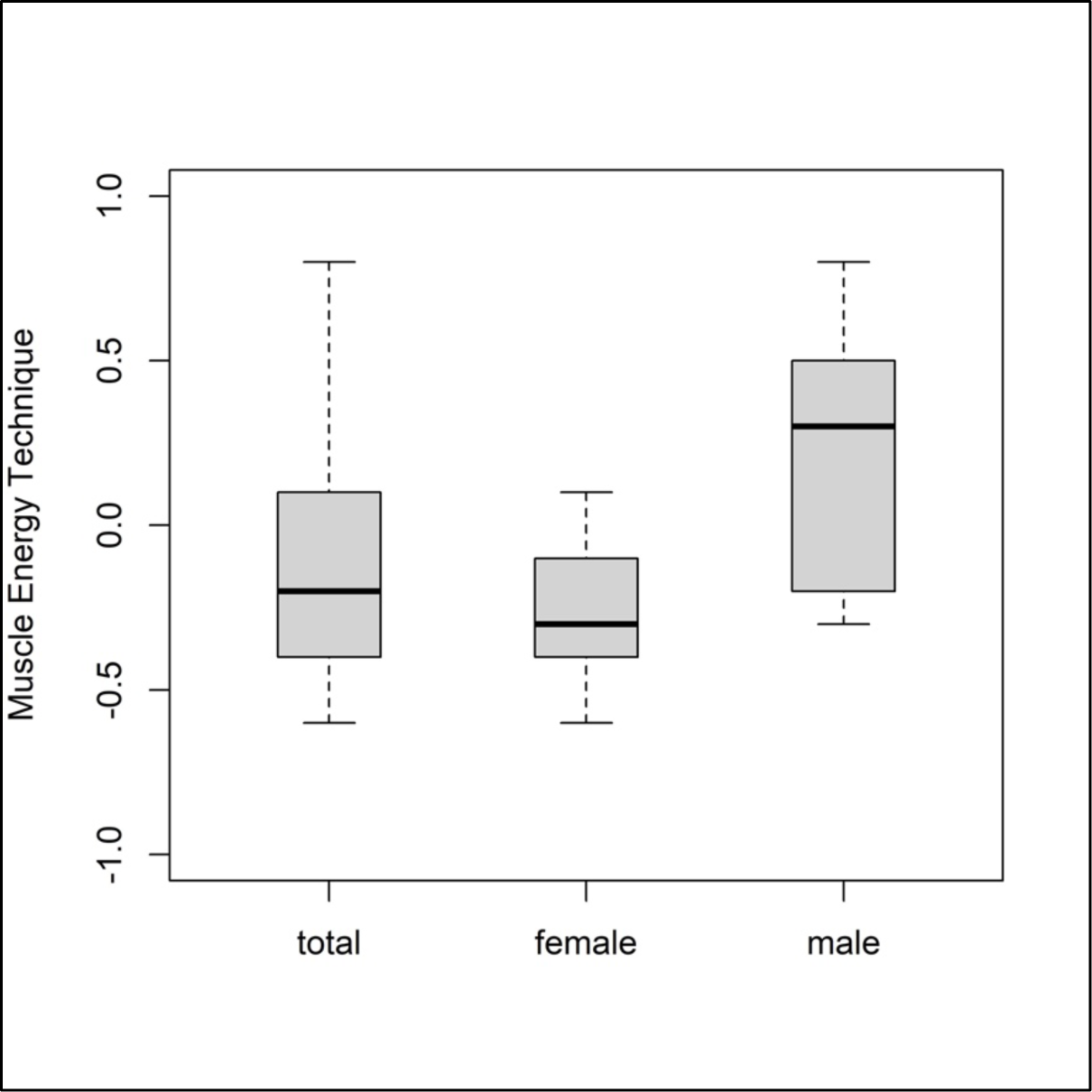
Mean differences in frequency of UT for MET; **Legend**: Not applicable.

**Figure 5.**
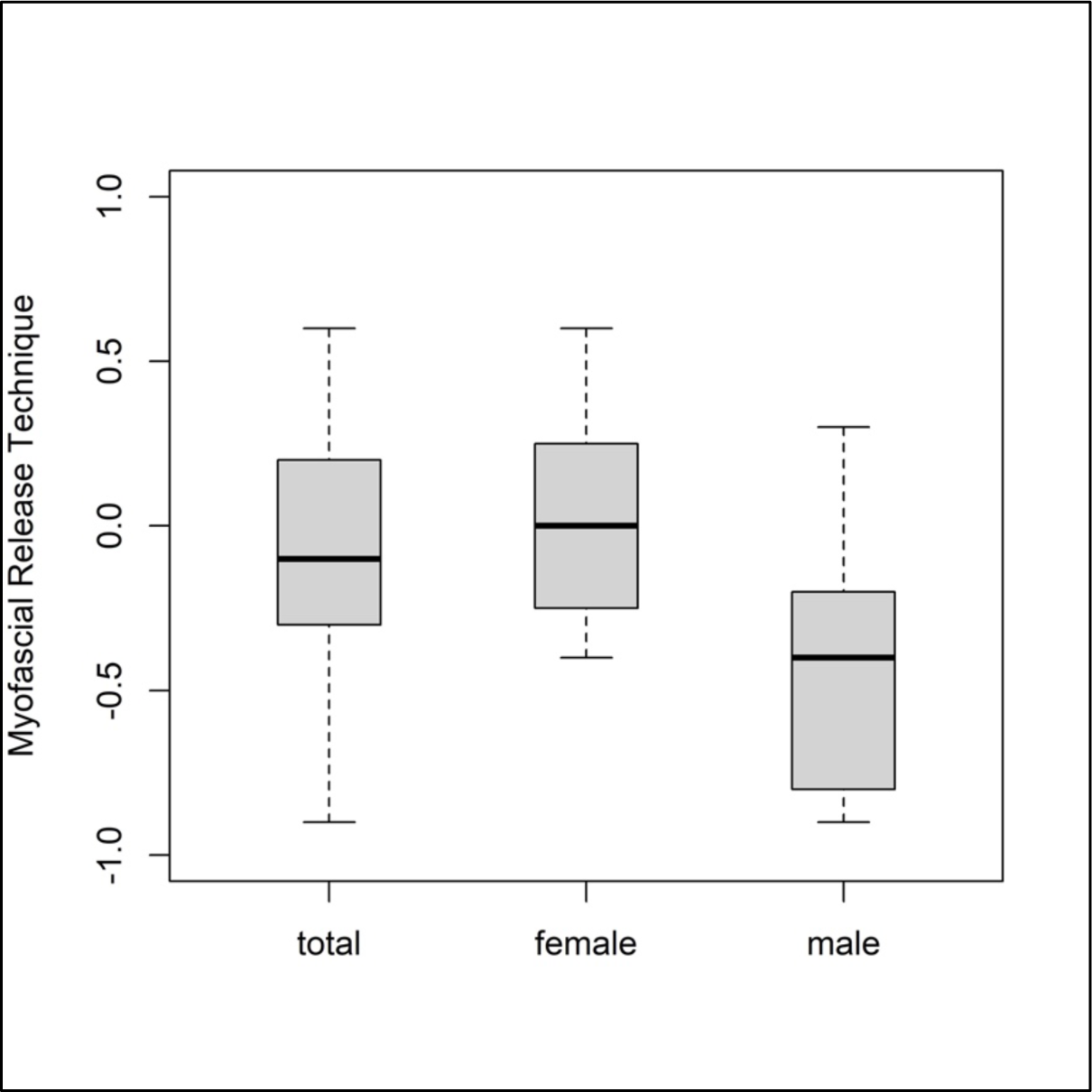
Mean differences in frequency of UT for MRT; **Legend**: Not applicable.

**Table 7.**
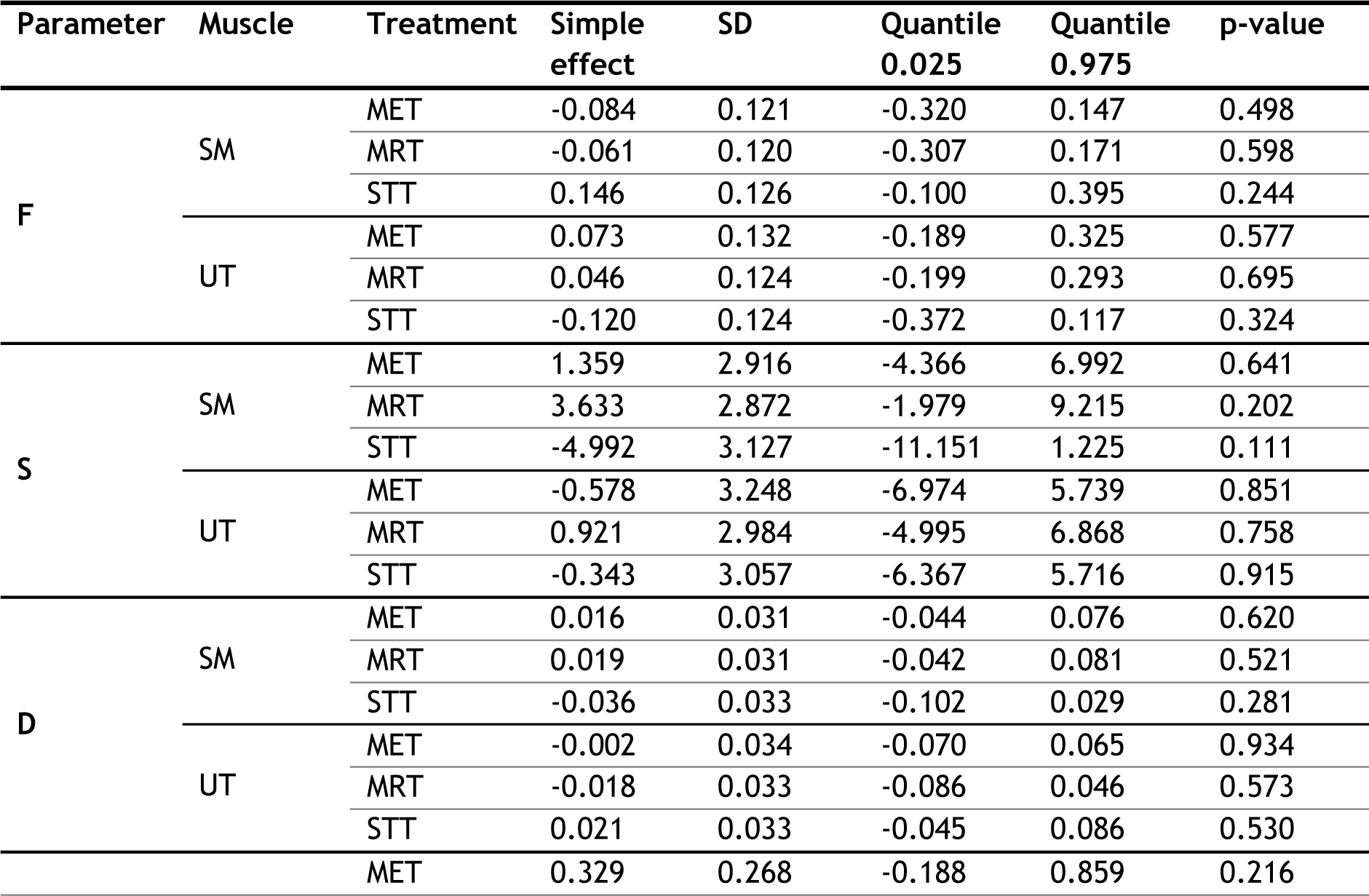
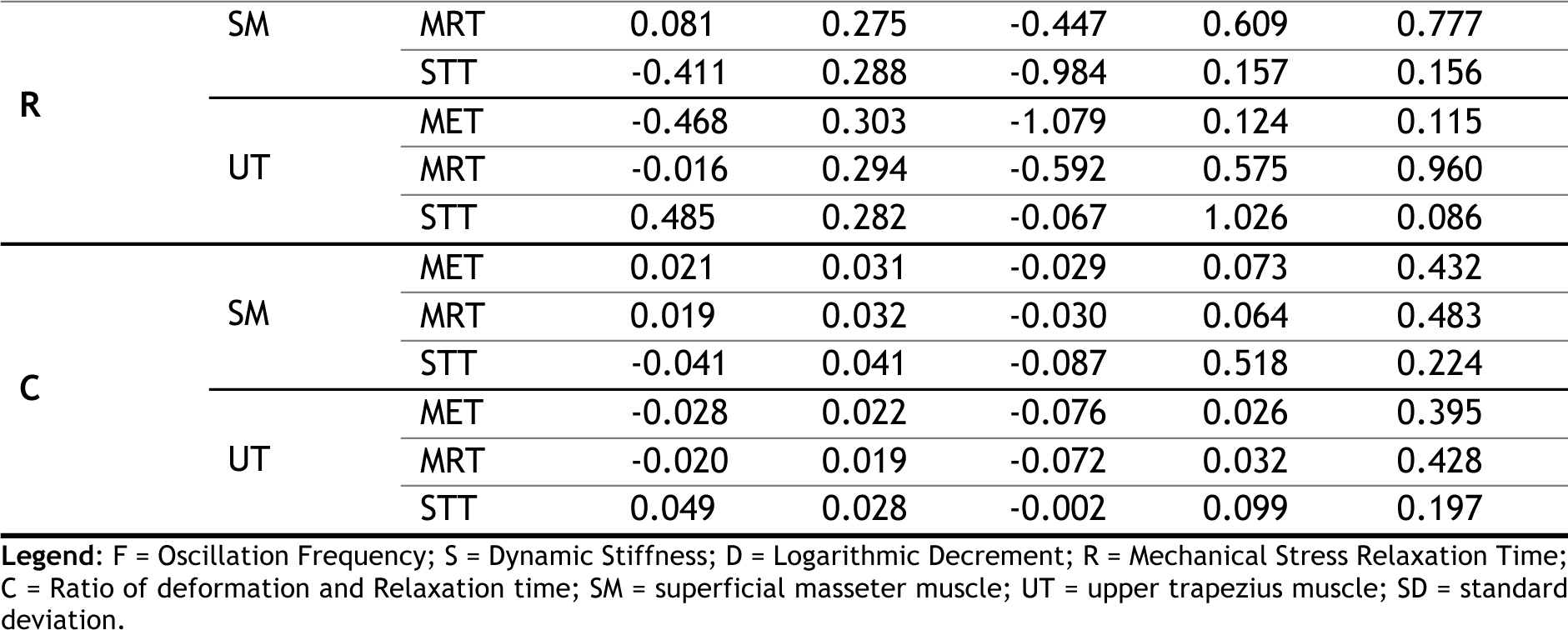
Results – Outcome for the secondary objective.

**Table 8.** Sex-specific analysis of frequency.

| Gender | Muscle | Treatment | Mean | Quantile | Quantile | p-value |
| --- | --- | --- | --- | --- | --- | --- |
|  |  |  |  | 0.025 | 0.075 |  |
| female | SM | MET | -0.203 | -0.526 | 0.117 | 0.999 |
|  |  | MRT | -0.417 | -0.771 | -0.064 | 0.189 |
|  |  | STT | -0.083 | -0.463 | 0.284 | 0.506 |
|  | UT | MET | -0.259 | -0.675 | 0.149 | 0.443 |
|  |  | MRT | 0.025 | -0.315 | 0.365 | 0.177 |
|  |  | STT | -0.274 | -0.620 | 0.069 | 0.637 |
| male | SM | MET | -0.369 | -0.915 | 0.168 | 0.276 |
|  |  | MRT | 0.101 | -0.404 | 0.604 | 0.199 |
|  |  | STT | 0.107 | -0.525 | 0.723 | 0.411 |
|  | UT | MET | 0.222 | -0.371 | 0.798 | 0.195 |
|  |  | MRT | -0.400 | -0.932 | 0.131 | 0.199 |
|  |  | STT | -0.296 | -0.826 | 0.248 | 0.407 |

Legend: SM = superficial masseter muscle; UT = upper trapezius muscle; SD = standard deviation. P-value for the simple effects.

### Harms

Participants were instructed to report harms to their physical or mental health to the principal investigator by phone or e–mail, pending their severity. No harms were reported.

## DISCUSSION

### Findings

To the best of our knowledge, this study is the first to investigate the immediate effect of OMT on the HRMT of healthy subjects. In detail, we used biomechanical and viscoelastic measures to assess the effect of manual techniques with different modalities on muscles with different characteristics. The sample comprised 57 participants (computed: 60; screened: 82; randomised 59) and showed acceptable recruitment and retention rates (72% and 96%, respectively). First, we report significant results for the primary objective (main effect). In detail, MET, MRT, and STT applied to the SM and UT (CS was excluded) resulted in significantly decreased muscle tone (F [p=0.008]), decreased biomechanical (S [p=0.048] not D [p=0.527]), and increased viscoelastic properties (R [p<0.001] and C [p=0.028]) from baseline to follow-up (compared to the same, but untreated, muscles of subjects from other groups) **(Table 6)**. Notably, decrement (D [p=0.527]) did not change significantly, however, this muscle parameter was previously proposed to be a constant [130]. The data suggest that the primary hypothesis can be confirmed, showing that osteopathic techniques modulated the HRMT of treated muscles in healthy subjects by decreasing muscle tone and stiffness while increasing relaxation and creep (decrement did not change significantly). Second, we demonstrate no significant results for the secondary objective (interaction effect). However, we detected a non-significant trend (p>0.05) suggesting that the decrease in muscle tone (F), decrease in biomechanical (D not S), and increase in viscoelastic properties (R and C) were achieved through MET and MRT (not STT) when applied to the SM, and through STT (not MET and MRT) when applied to the UT **(Table 7)**. Notably, this tendency was consistent for all muscle properties except for muscle stiffness (S). In other words, the smaller and thinner muscle (SM) responded (as expected) to the active (MET) and low-pressure technique (MRT), whereas the larger and thicker muscle (UT) responded (as expected) to the high-pressure technique (STT). The reported trend is fairly consistent with the secondary hypothesis (considering that the CS was excluded) but requires further scrutiny for verification or falsification. Lastly, subgroup analysis for the main effect revealed a significant sex-specific difference for muscle tone (p=0.030) but no other muscle properties (p>0.05). Overall, female subjects showed greater descriptive changes (mean values for primary objective) in all muscle properties than male subjects **(Table 6)**. For the interaction effect, no significant sex-specific difference was found but an interesting tendency showed that the muscle tone of the UT increased after MET and decreased after MRT in males, whereas the opposite was reported for females **(Table 8)** (**Figure4**) (**Figure 5**). The reason for this trend is unclear but we hypothesise, based on clinical experience, that males tend to exaggerate the counterpressure during MET techniques (presumably to demonstrate strength) which may have increased muscle tone.

### Mechanisms

To date, the mechanisms underlying these reported changes in HRMT following OMT are unclear and require further investigation. There might be biological, psychological, and social factors involved. However, based on the context of this study (comprising a short treatment time, strong manual focus, and unsound therapeutic alliance), we speculate that a biological mechanism of action is most probable. Though it might be the case that the three techniques have the same, different, overlapping, or multiple mechanisms of action **(Box 2)**, we suggest that mechanotransduction may underlie these changes. In detail, mechanotransduction suggests that extracellular mechanical signals are converted into intracellular chemical signals (and changes in gene expression) via integrins, which physically couple the extracellular matrix (including collagen fibres) with the intracellular cytoskeleton (including actomyosin filaments) [131]. As the HRMT depends on the interaction of cellular actomyosin filaments [79], we hypothesise that mechanical stimuli provided within osteopathic treatment may change the tensional forces within the (collagen fibres of the) extracellular matrix and, through integrins, within the (actomyosin filaments of the) intracellular cytoskeleton, thereby modifying the HRMT.

### Comparison

Overall, there is a scarcity of studies assessing the effect of manual treatment on muscle properties. Moreover, the available literature is focused on muscle stiffness and tone, whereas decrement, relaxation, and creep are mostly not considered. Therefore, our findings complement the existing body of evidence. Taking these factors into account, our results are largely consistent with the current literature. For example, in healthy participants, it was shown that manual therapy can reduce muscle stiffness [132]; this was demonstrated for MET [133], MRT (or self-MRT, respectively, which mimics manual MRT with a foam roller) [134], and STT (or massage, respectively, which resembles STT using deep pressure gliding strokes) [135]. Similarly, it was revealed that trigger points have an increased muscle tone and stiffness [113], which can be decreased through manual myofascial release [136]. Notably, manual techniques might also reduce muscle tone and stiffness in participants with MSDs [137]. Particularly in patients with LBP, increased paravertebral muscle tone and stiffness were demonstrated [138], which could be reduced through manual techniques like spinal mobilisation [139]. However, further research is required to substantiate these findings. Beyond that, we need to consider other factors that could have swayed the results, such as gender, age, and handedness. The sample in this study was relatively homogenous comprising predominantly female (68%), young (22.7 ± 4.5 years), and right-handed (91%) subjects. In general, muscle tone and stiffness are reported to be greater in males than females [140–142], however, there are also conflicting findings [105]. Nonetheless, higher muscle tone and stiffness in males may arise due to differences in the composition of muscle fiber types between the genders [143] as well as physiological varieties in muscle size, mass, and conditioning [130]. Besides gender, significant differences in muscle stiffness have also been reported between age groups (higher in elderly and middle-aged than young individuals) [142] and handedness (higher on the side of the dominant hand) [141]. Although subgroup analysis was performed for gender, we did not consider the impact of age (because only two subjects were over thirty years old) and handedness (because all measures were taken on the right side and only five subjects were left-handed).

### Limitations

There are limitations to consider when interpreting the results of this study. First and foremost, one of the three conditions was dropped because of frequent measurement errors. In detail, all missing values (8.9% of all numbers analysed) originated from myotonometer measurements of the CS (not SM or UT), which was consequently excluded from the analysis. These dependent missing values limited the number of observations available to analysis and therefore reduced the statistical power [144]. Further, dropping one condition increases the likelihood of reporting false-positive results [145]. Presumably, these missing values arose because the feasibility of the MyotonPRO is limited to measurements of muscles that are thicker than 3mm [111] and not located near the bone [146, 147] **(Box 6)**.

#### Box 6 CS characteristics

Prior to the trial, we reviewed the literature on the thickness of the CS, which was reported with a mean maximum thickness of 5.50 ± 0.91 mm in healthy subjects [94]. However, because the measurements of the CS were frequently inconsistent, we reviewed the literature again and found other studies accounting for an average thickness of approximately 1.62 ± 0.4 mm [97], 2-3 mm [148], and ∼2.4-2.8 mm [149], respectively. Thus, we acknowledge a flaw in our initial literature search and suggest that the CS may be too thin and/or near the bone to be consistently measured using the MyotonPRO.

Another shortfall is that the results are merely informative of an immediate-term effect (approximately five minutes after treatment). Thus, the measured tissue response may be reflective of the thixotropy effect [150]; albeit the size of this effect remains unclear [151]. Further, it is noteworthy that participants were assessed in supine position although the HRMT relates to the biotensegrity system and posture [79]. Other barriers to interpreting the results relate to the interventions. For one, manual techniques are often loosely defined, and it was particularly difficult to find literature describing the same manual procedure as the STT used in this study. In the end, we settled to include research on massage techniques, which employ similar parameters than the STT. Future research might consider not examining manual techniques (e.g., MET, MRT, and STT) but rather their biophysical parameters (e.g., stretch, compression, shear, and torque forces) to ensure optimal comparability [152, 153]. Furthermore, the manual techniques were applied for five minutes, which appears brief but seems to be sufficient for one muscle to be treated with one technique (because the therapists perceived palpatory signs of release/relaxation). Also, we cannot rule out therapist-specific differences in outcomes, although consensus training was implemented. Moreover, the interventions encompassed common manual techniques with different characteristics, however, single manual techniques are not representative of person-centered osteopathic care.

### Future directions

Future studies might assess the effect of OMT on the HRMT of people with MSDs because conditions like LBP and NP are associated with altered muscle properties [116, 138, 154]. For example, in patients with LBP compared to healthy controls, the lumbar extensor myofascia generally shows increased muscle tone (F), stiffness (S), and decrement (D) [141, 154–156] as well as decreased relaxation (R) and creep (C) [138, 157, 158]. Hence, we propose to examine the effect of OMT on the muscle properties of, for example, the lumbar extensor myofascia or the upper trapezius muscles in patients with LBP or NP, respectively. Based on the present findings, we hypothesise that osteopathic interventions will decrease muscle tone, stiffness, and decrement, and increase relaxation and creep in this population. However, future studies are needed to validate or falsify this hypothesis. Another consideration for future research is that muscle properties like stiffness do not seem to correlate with pain (in patients with chronic NP and LBP), because muscle stiffness typically returns to baseline one day after treatment (using cupping massage) even if the pain improves [159]. Lastly, it is unclear if OMT could also have the opposite effect. In detail, it has previously been assumed that OMT restores normal muscle tone where it is altered [73–76], meaning that high muscle tone decreases, and low muscle tone increases, when OMT is applied [83]. Therefore, it might be useful to assess the effect of OMT on conditions associated with both hypertonia and hypotonia. Lastly, future research should assess if these changes are enduring in the short, medium, and long term.

### Interpretation

In this study, we demonstrate that OMT modifies the HRMT in healthy participants without significant interaction of muscle-technique pairs. The mechanisms underlying these changes are unclear and the results are limited by the exclusion of one condition. Our findings are largely consistent with previous research but merely informative of an immediate effect. Future studies should modify the protocol and assess if these effects are reproducible (and beneficial) in patients with MSDs. In the end, although speculative, we hypothesise that modifying the HRMT may be a mechanism of action underlying manual techniques.

## CONCLUSION

Taken together, we built on research suggesting that alterations of the HRMT may underlie MSDs and respond to OMT. Taken together, we report that osteopathic techniques (MRT, MET, and STT) significantly decreased muscle tone (F), decreased biomechanical (S not D), and increased viscoelastic properties (R and C) of the SM and UT (CS was not measurable) at immediate term. Hence, osteopathic modalities seem to modify the HRMT in healthy subjects, generating changes likely associated with health benefits (decreasing muscle tone and stiffness while increasing viscoelasticity). However, it is unclear if some muscle–technique pairs change the HRMT more effectively than others (e.g., techniques using high pressure applied to thicker, rather than thinner, muscles) and future studies are required to answer this inquiry. More importantly, these findings may inform future research assessing the effect of OMT on the HRMT in patients with MSDs.

## Data Availability

All data produced in the present work are contained in the manuscript.

## Abbreviations

MRT: Myofascial Release Technique
MET: Muscle Energy Technique
STT: Soft Tissue Technique
CS: Corrugator Supercilii Muscle
SM: Superficial Masseter Muscle
UT: Upper Trapezius Muscle
F: Frequency
S: Dynamic Stiffness
D: Logarithmic Decrement
R: Mechanical Stress Relaxation Time
C: Ratio of Deformation and Relaxation Time
MSDs: Musculoskeletal Disorders
HRMT: Human Resting Muscle Tone
OMT: Osteopathic Manipulative Treatment
EMG: Electromyography
MPs: Measurement Points

## Acknowledgements

We like to express our gratitude to **(i)** the students and staff of the Osteopathie Schule Deutschland for their participation and assistance during the trial; **(ii)** Alexander Karner, MSc, Christian Selzle, MSc, Stephan Rohde, MSc, and Charline Krohn for providing intellectual, organizational, and spatial support; **(iii)** Ass. Prof. Jan Porthun, MMMSc for helpful discussions about the study design and statistics; and **(iv)** Dr. Christine Lohr, MSc, and Marius Dickmann, MSc for proofreading the manuscript and providing thoughtful comments.

## Authors’ contributions

**LB**: Conceptualization, Methodology, Writing – Original Draft, Writing – Review & Editing, Visualization, Supervision, Project administration; **JS**: Formal analysis, Investigation, Writing – Review & Editing, Visualization; **JR**: Investigation, Writing – Review & Editing; **BG**: Investigation, Writing – Review & Editing, Project administration; **CL**: Investigation, Writing – Review & Editing; **AK**: Formal analysis, Writing – Review & Editing; **AB**: Formal analysis, Writing – Review & Editing; **TS**: Writing – Review & Editing, Supervision.

## Availability of data and materials

The datasets used and/or analysed during the current study are available from the corresponding author on reasonable request.

## Ethics approval and consent to participate

This study has obtained informed consent from participants, was carried out in accordance with the Declaration of Helsinki and was approved by the ethics committee of the Osteopathic Research Institute (Nr. 020-01).

## Consent for publication

Written informed consent for publication of the images was obtained from the subjects.

